# Differences in sleep spindle wave density between patients with diabetes mellitus and matched controls: implications for sensing and regulation of peripheral blood glucose

**DOI:** 10.1101/2024.04.11.24305676

**Authors:** Deryck Yeung, Amlan Talukder, Min Shi, David M. Umbach, Yuanyuan Li, Alison Motsinger-Reif, Zheng Fan, Leping Li

## Abstract

**Background:** Brain waves during sleep are involved in sensing and regulating peripheral glucose level. Whether brain waves in patients with diabetes differ from those of healthy subjects is unknown. We examined the hypothesis that patients with diabetes have reduced sleep spindle waves, a form of brain wave implicated in periphery glucose regulation during sleep.

**Methods:** From a retrospective analysis of polysomnography (PSG) studies on patients who underwent sleep apnea evaluation, we identified 1,214 studies of patients with diabetes mellitus (>66% type 2) and included a sex- and age-matched control subject for each within the scope of our analysis. We similarly identified 376 patients with prediabetes and their matched controls. We extracted spindle characteristics from artifact-removed PSG electroencephalograms and other patient data from records. We used rank-based statistical methods to test hypotheses. We validated our finding on an external PSG dataset.

**Results:** Patients with diabetes mellitus exhibited on average about half the spindle density (median=0.38 spindles/min) during sleep as their matched control subjects (median=0.70 spindles/min) (*P*<2.2e-16). Compared to controls, spindle loss was more pronounced in female patients than in male patients in the frontal regions of the brain (*P=*0.04). Patients with prediabetes also exhibited signs of lower spindle density compared to matched controls (*P=*0.01-0.04).

**Conclusions:** Patients with diabetes have fewer spindle waves that are implicated in glucose regulation than matched controls during sleep. Besides offering a possible explanation for neurological complications from diabetes, our findings open the possibility that reversing/reducing spindle loss could improve the overall health of patients with diabetes mellitus.

**Funding:** This research was supported by the Intramural Research Program of the National Institutes of Health, National Institute of Environmental Health Sciences (ZIA ES101765).

## INTRODUCTION

Diabetes mellitus is a major public health concern. In the United States, the Centers for Disease Control and Prevention (CDC) estimates that 38.4 million people (11.6% of the US population) suffer from diabetes and an additional 97.6 million people aged 18 years or older (38.0% of the adult US population) have prediabetes. Globally, an estimated 537 million (10.5%) adults aged 20–79 years are currently living with diabetes and the number is expected to rise to 783 million (12.2%) by 2035 (***edition 2021***).

Diabetes mellitus is characterized by metabolic dysregulation resulting from impaired insulin secretion, insulin resistance, or a combination of both (***Bastaki 2019; DeFronzo et al. 2015; Katsarou et al. 2017; Olokoba et al. 2012; Reed et al. 2021***). Among all diabetes, type 2 diabetes mellitus accounts for more than 90% of all cases. Diabetes mellitus is associated with several complications including coronary artery disease, chronic kidney disease, neuropathy, cancer, obstructive sleep apnea, and liver disease (***Collaborators 2023; Tomic et al. 2022; van Dieren et al. 2010***). Diabetes is also associated with neurological conditions including cognitive impairment and memory loss (***Biessels and Despa 2018; Kodl and Seaquist 2008***), dementia (***Barbiellini Amidei et al. 2021; Biessels et al. 2006; Biessels and Despa 2018***), and Alzheimer’s disease (***Ahtiluoto et al. 2010; Antal et al. 2022; Arnold et al. 2018; Kellar and Craft 2020; Moreno-Gonzalez et al. 2017; Ott et al. 1999; Sims-Robinson et al. 2010***). Diabetes is manifested in the brain through neuronal death (***Moheet et al. 2015; Seaquist 2010***).

Neurological complications in patients with diabetes mellitus may be evident in their brain wave oscillations measured by electroencephalograms (EEGs). Selective suppression of slow waves in non-rapid eye movement (NREM) sleep in healthy subjects is associated with the risk for type 2 diabetes in humans (***Johnson et al. 2022; Tasali et al. 2008***); however, little is known about changes in brain waves in patients with diabetes mellitus during sleep.

The brain waves during sleep play a recently recognized but important role in regulating peripheral glucose metabolism. The neocortex, hippocampus, and thalamus interact through coupling of distinct EEG oscillations during sleep: slow waves (0.5-1.5Hz), spindles (10-15Hz), and sharp wave-ripples (∼80–100 Hz in human); this interaction is associated with important physiological functions such as transferring, exchanging, and consolidating neuronal information during sleep (***Buzsáki 2015; Cox et al. 2020; Eschenko et al. 2008; Geva-Sagiv et al. 2023; Helfrich et al. 2019; Joechner et al. 2023; Karimi Abadchi et al. 2020; Mölle et al. 2006; Ngo et al. 2020; Oyanedel et al. 2020; Siapas and Wilson 1998; Staresina et al. 2023***). Recently, it has been demonstrated that hippocampal slow-wave sharp ripples also play a role in modulating peripheral glucose homeostasis (***Tingley et al. 2021***). Clustered events of slow-wave sharp ripples were found to be correlated with reduced periphery glucose level. Similarly, Vallat et al. showed that coupling between slow-wave oscillations and spindles during sleep predicts next-day insulin dependent glucose regulation (***R. Vallat et al. 2023***). Suppression of slow-wave sleep in healthy subjects also resulted in marked decreases in insulin sensitivity (***Tasali et al. 2008***). Taken together, those findings strongly suggest that brain waves are implicated in peripheral glucose regulation, further supporting the bidirectional relationship between diabetes and sleep (***Briancon-Marjollet et al. 2015; Cappuccio et al. 2010; Chen et al. 2023; Farabi et al. 2016; Noga et al. 2024; Schipper et al. 2021; Spiegel et al. 1999***).

Spindle wave, also called sleep spindle, is one of the brain waves. Sleep spindles are primarily generated by the thalamus and are an EEG hallmark of the NREM sleep (***Andrillon et al. 2011; Fernandez and Lüthi 2020; Peyrache and Seibt 2020; Purcell et al. 2017; Schönauer and Pöhlchen 2018***). Spindles are believed to mediate many sleep-related functions, including memory consolidation and cortical development (***Andrillon et al. 2011; Fernandez and Lüthi 2020; Peyrache and Seibt 2020; Purcell et al. 2017***). Dysregulation of spindles is associated with neurodegenerative disorders (***Ferrarelli et al. 2007***) and aging (***Purcell et al. 2017***). Accordingly, changes in spindle characteristics may be viewed as a marker for altered brain function.

The brains of patients with diabetes mellitus may have an impaired capacity for sensing and regulating peripheral glucose compared to those of healthy subjects. We hypothesize that the generation of spindles during the NREM sleep is lower in patients with diabetes mellitus, disrupting the slow wave−spindles−sharp wave ripple regulatory network and resulting in impaired sensing and regulation of peripheral glucose levels. In this study, we used EEG data from in-laboratory polysomnography (PSG) studies to compare six spindle characteristics including amplitude, relative power, density, oscillation, frequency, and symmetry between 1,214 patients with diabetes and controls subjects matched individually on age and sex. We found that patients with diabetes mellitus had significantly lower spindle density compared to matched control subjects. We validated our findings on an external PSG dataset from the Sleep Heart Health Study (SHHS) (***Quan et al. 1997; Zhang et al. 2018***).

## Materials and Methods

### Subjects and study protocol

This study involved retrospective review of patient records. We downloaded de-identified records of patients referred by their health-care providers for PSG studies at the sleep laboratory at the University of North Carolina at Chapel Hill (UNC-CH) between January 2019 and March 2023. This sleep laboratory is accredited by the *American Academy of Sleep Medicine* (AASM). Reasons for patient referral were not included in our data, though most were likely referred for evaluation of sleep apnea.

From the records of 10,707 PSG studies downloaded, we identified 1,897 PSG studies from 1,474 patients who were 18 yrs or older with diabetes mellitus. When we identified multiple PSG studies for any single patient, we used the one with the youngest age. We further removed those with fewer than 100 minutes of total sleep time (n=108). This resulted in 1,214 PSG studies from 1,214 diabetes patients (26 with type 1, 799 with type 2, and 389 unspecified). The patient inclusion flowchart is shown in Supplemental Figure 1.

**Figure 1.**
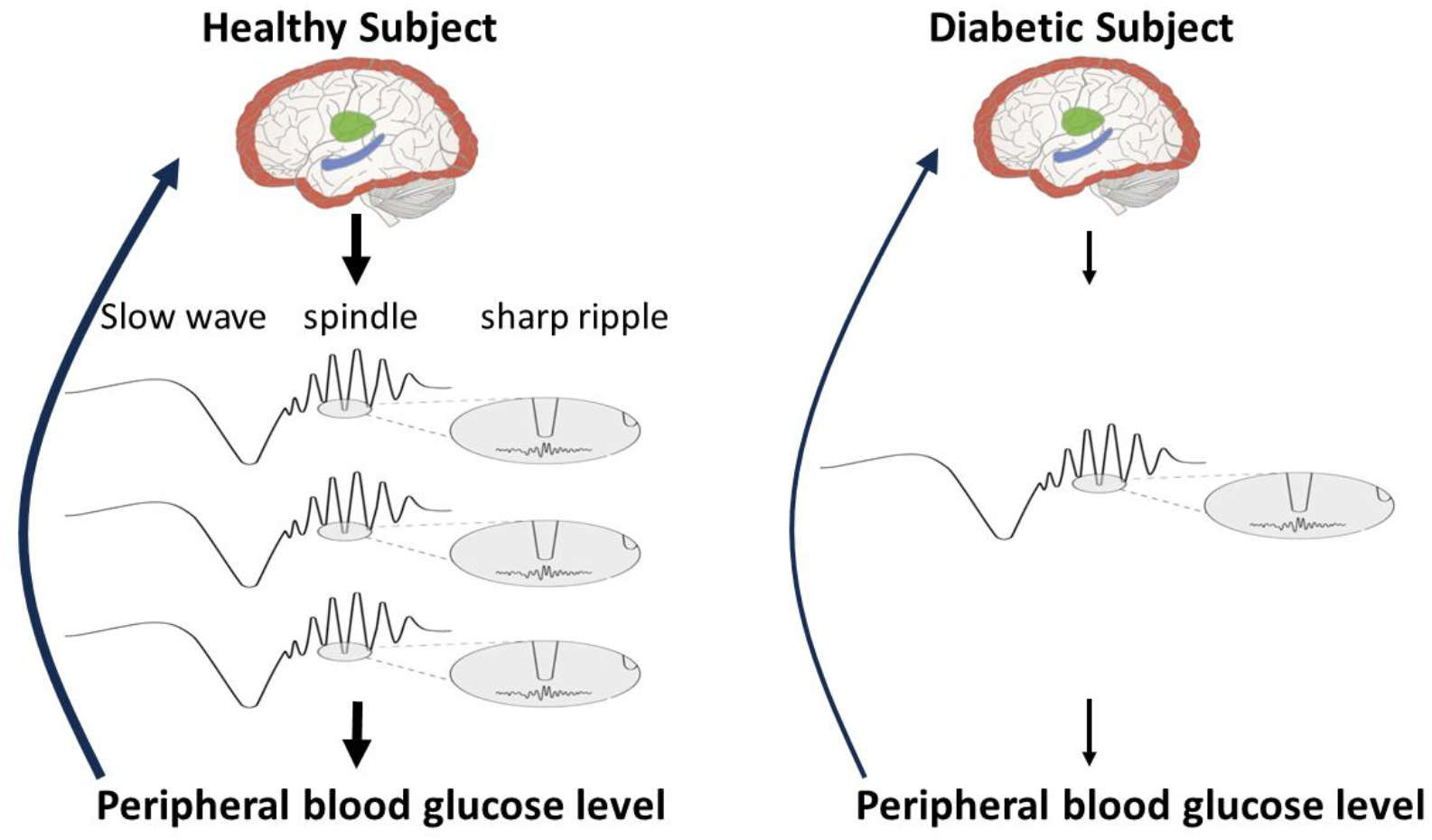
Schematic of brain wave regulation of peripheral blood glucose level. Arrow thickness is proportional to the strength of presumed response. Left: thalamic-hippocampal-neocortical interaction via three neural oscillations – slow waves (∼ 1Hz) from neocortex (red), spindles (∼10-15Hz) from thalamus (green) and sharp wave ripples (∼100Hz) from hippocampus (blue) – senses and regulates peripheral blood glucose level during sleep. Right: our hypothesis – patients with diabetes have fewer thalamocortical spindles, impairing brain sensing and regulation of peripheral blood glucose level.

To select PSG studies for control subjects matched to those from diabetes patients, we followed the same procedure reported previously (***Talukder et al. 2024***). Briefly, we excluded all PSG studies for which the subjects were diagnosed with major medical disorders such as cancer, diabetes, major depression, brain and cardiopulmonary diseases. If a subject had multiple studies, retained the single study with the youngest age. For each diabetes study, we randomly selected a control study of the same sex and similar age (± 1 yr) as the matched control (Table 1).

**Table 1.** Age and sex distribution of patients with diabetes and their age- and sex-matched control subjects.

| Data | Sex | Group | N | Min. | 1<sup>st</sup> Qu. | Median | Mean | 3<sup>rd</sup> Qu. | Max. |
| --- | --- | --- | --- | --- | --- | --- | --- | --- | --- |
| Our<br>PSG | Male | Diabetes | 524 | 18.0 | 52.2 | 60.5 | 60.3 | 69.9 | 88.3 |
|  |  | Control |  | 18.3 | 52.3 | 60.7 | 60.2 | 69.8 | 88.5 |
|  |  | Diabetes-Control |  | -1.0 | -0.5 | 0 | 0.1 | 0.6 | 1.0 |
|  | Female | Diabetes | 690 | 18.1 | 50.6 | 59.1 | 58.6 | 67.0 | 89.7 |
|  |  | Control |  | 17.1 | 51.0 | 59.0 | 58.6 | 67.2 | 89.1 |
|  |  | Diabetes-Control |  | -1.0 | -0.5 | 0.0 | 0.0 | 0.5 | 1.0 |
| SHHS<br>PSG | Male | Diabetes | 233 | 43.0 | 64.0 | 70.0 | 69.3 | 76.0 | 89.0 |
|  |  | Control |  | 44.0 | 63.0 | 70.0 | 69.2 | 75.0 | 90.0 |
|  |  | Diabetes-Control |  | -1.0 | -1.0 | 0 | 0 | 0 | 1.0 |
|  | Female | Diabetes | 182 | 40.0 | 62.0 | 71.0 | 69.0 | 77.0 | 90.0 |
|  |  | Control |  | 41.0 | 62.0 | 71.5 | 68.9 | 76.8 | 89.0 |
|  |  | Diabetes-Control |  | -1.0 | -1.0 | 0 | 0.1 | 1.0 | 1.0 |

Similarly, we identified 376 PSG studies for 376 patients with prediabetes; none were among the diabetes patients. We applied the same procedure and criteria to select matched controls for prediabetes subjects as we did for diabetes patients except that subjects with prediabetes were now excluded when selecting control subjects. 214 control subjects were matched to both a diabetes and a prediabetes patient.

This study was approved by the Institutional Review Board of UNC-CH (IRB #21-1984).

### In-laboratory PSG study

Each study included an electroencephalogram with at least six channels (frontal F3 and F4, central C3 and C4, and occipital O1 and O2, plus two reference channels M1 and M2). The multi-channel PSGs were recorded digitally and stored using a *Natus* polygraph (Natus® NeuroWorks®). Studies were manually scored following guidelines from the AASM scoring manuals and interpreted by physicians certified by the *American Board of Sleep Medicine*. Sleep staging/scoring to determine aspects of sleep architecture were carried out manually by AASM certified supervisory sleep technicians, and further verified by AASM-certified reading physicians from video, using guidelines current at the time of the PSG. The variables extracted include total sleep time, sleep latency, sleep efficiency, and duration of stage 2 (N2) of the NREM sleep.

### External dataset for validation

We validated our finding on an external dataset from the SHHS study which was designed to determine the cardiovascular and other consequences of sleep-disordered breathing (***Quan et al. 1997; Zhang et al. 2018)***. This dataset contains 5,804 samples (2,765 males and 3,039 females) aged from 39 to 90 yrs. Among the 5,804 participants, we found EEG data for 405 subjects with self-reported diabetes. One subject did not have enough sleep time and was removed from further analysis. To select for controls, we excluded those with major medical conditions such as stroke, chronic obstructive pulmonary disease, and heart disease. For each of the 404 subjects with self-reported diabetes, we randomly selected a control subject of the same sex and similar age (± 1 yr) as the matched control (Table 1).

### EEG data processing

We extracted six channel EEG data and their corresponding sleep (N1, N2, N3, and R) and wake (W) from the *EDF* files using the Python *MNE* package (***Gramfort et al. 2013***), where N1, N2, and N3 are stages 1, 2, and 3 of the NREM sleep and R is the rapid-eye movement (REM) sleep. Each channel was re-referenced with respect to its opposite mastoid channel M1 or M2 (F3 -M2, F4-M1, C3-M2, C4-M1, O1-M2, and O2-M1). To each channel, we applied a zero-phase notch filter to remove a power-line noise at 60Hz. We removed epochs (30 seconds) with excessive flat signals (***Talukder et al. 2024***) as well as epochs with a signal amplitude greater than 2,000 µV. After these pre-processing steps, we carried out multitaper spectrogram analysis (***Prerau et al. 2017***) to calculate EEG power in the frequency range of 0.25 to 30Hz. We then removed epochs whose total power was beyond either the 1^st^ or the 3^rd^ quartile by more than three times the interquartile range (***Talukder et al. 2024***).

### Spindle analysis

We used the YASA software (***Raphael Vallat and Walker 2021***) to extract the following spindle characteristics: durations, amplitudes, absolute power, relative power, frequency, oscillations, and symmetry (***Purcell et al. 2017***) (Supplemental Figure 2). Spindle density is defined as the total number of spindles observed during the N2 stage of NREM sleep divided by the time spent in N2 sleep (e.g., minutes). For an individual spindle (Supplemental Figure 2), spindle frequency is defined as the number of cycles per second. Spindle amplitude is the difference between the maximum and minimum deviation (i.e., peak-to-peak) in voltage. Spindle power is the signal power in [11 Hz,15 Hz]. Relative spindle power is the spindle power divided by the total power in the frequency range from 1Hz to 30Hz. Number of oscillations is the number of cycles in a spindle. Spindle symmetry is based on the relative location of the spindle’s central “peak” (the point of maximum peak-to-peak amplitude) (***Purcell et al. 2017***). A subject-specific value of each characteristic is calculated by averaging spindle-specific values across all spindles that occurred during N2 sleep for the individual.

### Statistical analysis

To identify features that differed between patients with diabetes mellitus and their matched controls, we used two-sided Wilcoxon signed-rank tests applied to the patient-minus-control differences. To assess a possible trend in the matched patient-minus-control difference with age, we used simple linear regression with the difference as the response and the average age between patient and control as the predictor. To evaluate whether patient-minus-control differences depended on dichotomous factors that differed between matched pairs (such as sex or presence of complications in the patient), we used two - sided Wilcoxon rank-sum tests unless specified otherwise. For statistical analyses of sleep architecture variables, we restricted attention to diagnostic PSG studies. We regarded P values <0.05 as statistically significant.

## RESULTS

### Characteristics of patients with diabetes and their matched control subjects

Among the 1,214 patients older than 18 years of age with diabetes mellitus, 524 (46.2%) were males and 690 (53.8%) were females (Table 1). The median ages for male patients and their matched controls were 60.5 and 60.3 yrs, respectively, and the median difference in age between paired male subjects (patient minus control) was 0 yrs. For women, the corresponding median ages were 59.1 and 58.6 yrs, respectively, with a median difference between paired subjects of 0 yrs.

Among those with diabetes mellitus, 257 (21.1%) were diagnosed with coronary artery disease (CAD), 130 (10.7%) with chronic kidney disease (CKD), 217 (17.8%) with neuropathy, and 61 (5%) with stroke (Supplemental Table 1). More than half of the patients (711, 58.3%) did not have any of the above four complications. In comparison, small percentages (0.3-6.6%) of the control subjects had any one of the four complications and most (89.4%) had none of them (Supplemental Table 1).

Nearly all (1,064, 87.6%) of the patients with diabetes mellitus had a prescription for one of the several classes of diabetes medications (Supplemental Table 1). The prescriptions most widely used among our sample of patients included: metformin (biguanide class) by 689 (56.7%) patients, various forms of insulin by 629 (51.8%), and GLP-1 (glucagon-like peptide 1) agonists by 306 (25.2%). Interestingly, 57 (4.6%) of the controls had prescriptions for metformin, though only small percentages (0.3-1.2%) of the controls had prescriptions for other diabetes medications. None had an insulin prescription.

### Comparison of sleep architecture

Both the patients with diabetes mellitus and matched controls took a similar amount of time to fall asleep (median=13.3 min compared to 14.4 min among controls) (*P=*0.9, Wilcoxon signed-rank test) (Supplemental Table 2). Patients with diabetes mellitus had shorter total sleep time compared to the matched controls (median=357.5 min compared to 365.0 min, *P=*0.009, Wilcoxon signed-rank test) (Supplemental Table 2). The median sleep efficiency among diabetes patients (79.2%) was slightly lower than that (80.2%) of the controls (*P=*0.08, Wilcoxon signed-rank test). Patients with diabetes mellitus had similar amount of time in N2 sleep (median=105.5 min) compared to the controls (median=107 min) (*P=*0.5, Wilcoxon signed-rank test) (Supplemental Table 2). For the matched diabetes/control pairs undergoing diagnostic PSG studies, patients with diabetes mellitus had overall higher sleep apnea/hypopnea index (AHI) (median=11.2 events/hr) compared to the controls (median=9.2 events/hr) (*P=*0.009, Wilcoxon signed-rank test) (Supplemental Table 2).

**Table 2.**
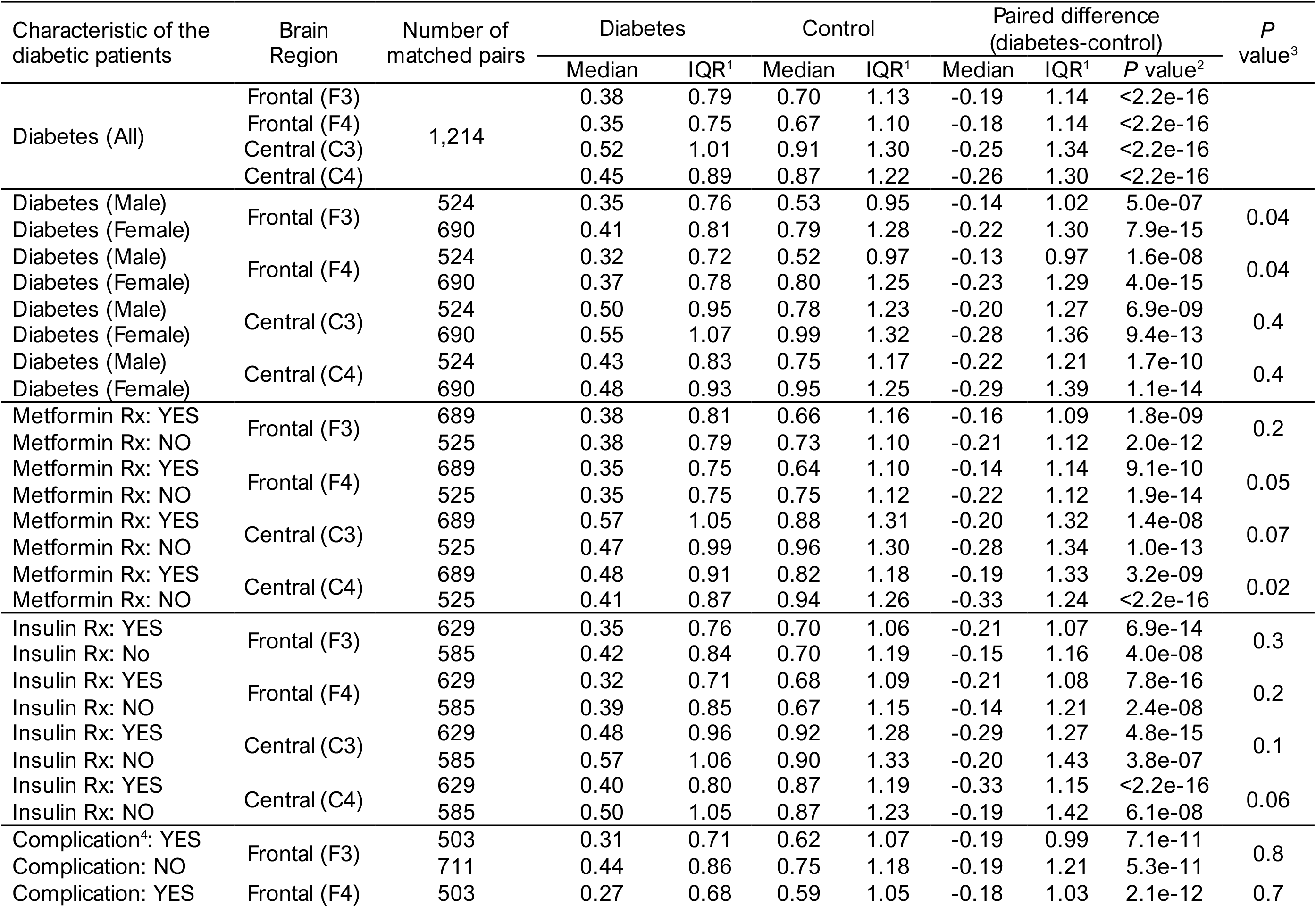
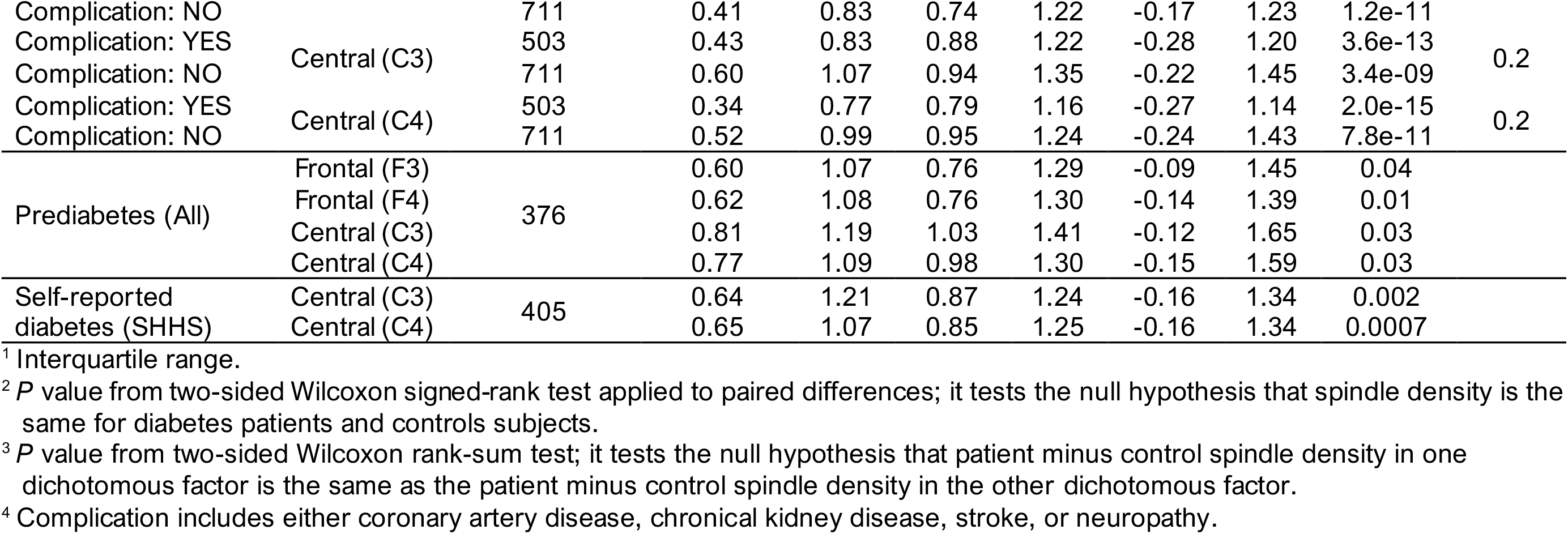
Comparison of spindle density between different subsets of patients with diabetes or with prediabetes and their age - and sex- matched control subjects.

### Comparison of spindle characteristics

We examined seven characteristics of spindles that occur during the N2 stage of NREM sleep: duration, amplitude, absolute power, relative power, frequency, number of oscillations, and symmetry (Supplemental Figure 2). Among the seven characteristics that we looked at, only spindle density was significantly different between patients with diabetes mellitus and matched controls (Supplemental Table 3). In all four regions (F3, F4, C3, and C4) of the brain, patients with diabetes mellitus had significantly lower spindle density compared to matched controls (Table 2, Supplemental Figure 3). For both diabetes patients and controls, spindle density was higher in the central regions (C3 and C4) than the frontal regions (F3 and F4) (*P* <2.2e-16, one-sided Wilcoxon signed-rank test, e.g., C3 > F3 or C4 > F4 for either diabetes or control). For each region of the brain, the median spindle density in patients with diabetes mellitus was nearly half of that in matched controls (Table 2). For a healthy subject with eight hours of sleep, 50% (4 hours) of total sleep time are typically in N2 stage of the NREM sleep. With a spindle density of 0.7 spindles/min (e.g., F3 in the control), for a healthy subject, there would be 168 spindles during a typical night of sleep. In comparison, patients with diabetes mellitus would produce just 91 spindles in the same region of the brain under similar sleeping conditions (F3, spindle density=0.38 spindles/min).

### Possible modifiers of patient-versus-control differences in spindle density

Our finding about differences in spindle density between diabetes patients and control subjects prompted us to examine some factors that might modify the observed association. We looked at four factors as possible modifiers: having a metformin prescription, presence of diabetes complications, sex, and age.

#### Prescriptions

Our sample included 678 diabetes patients who had a prescription for metformin and 525 without one; accordingly, we created two groups of matched pairs. In each group separately, patients with diabetes had significantly lower spindle density compared to matched controls in all four brain regions (Table 2). In addition, when we compared the magnitude of the difference between those two groups by brain region, we found the size of the differences (diabetes minus control) were lower among pairs where the patient had a metformin prescription (median diabetes minus control differences −0.16, −0.14, −0.20, and −0.19) compared to those without a metformin prescription (median diabetes minus control differences −0.21, −0.22, −0.28, and −0.33) (*P=*0.2, 0.05, 0.07, and 0.02 by Wilcoxon rank-sum test in brain regions F3, F4, C3, and C4, respectively), suggesting that patients with a metformin prescription had slightly better spindle density than those without a metformin prescription, especially in the right hemisphere of the brain (F4 and C4) (Table 2).

Our sample also included 629 diabetes patients with an insulin prescription and 585 without one. In each group separately, patients with diabetes had significantly lower spindle density than matched controls in all four brain regions (Table 2). The size of the difference in spindle density was slightly, but not significantly, larger in pairs where the patient had an insulin prescription (median diabetes minus control difference −0.21, −0.21, −0.29, and −0.33) compared to (median diabetes minus control differences −0.15, −0.14, −0.20, and −0.19) (*P=*0.3, 0.2, 0.1, and 0.06, for F3, F4, C3, and C4, respectively).

#### Diabetes complications

Our sample included 503 diabetes patients with at least one of the four major complications (CAD, CKD, stroke, and neuropathy) and 711 patients with none of them. Diabetes patients in each group showed significantly lower spindle density compared to matched controls in all four brain regions (Table 2). Comparison of the patient-minus-control difference between the two groups suggested that those without complications had slightly smaller differences in all four brain regions, though the comparison was not statistically significant in any region (*P=*0.8, 0.7, 0.2, and 0.2, respectively, for F3, F4, C3 and C4 was 0.30, Wilcoxon rank-sum test).

*Sex*. Our sample included 524 males and 690 females, each with their matched control subject. Both sexes showed significantly lower spindle density in diabetes patients compared to controls in all four regions of the brain (Table 2). We saw some evidence that the decline in spindle density in diabetes patients compared to controls was more profound in females compared to males, especially in the frontal regions of the brain (F3 and F4). The median differences between patients with diabetes and controls were −0.14 spindles/min in males and −0.22 spindles/min females in the left frontal region (F3) (*P=*0.04, Wilcoxon rank-sum test) and −0.13 in males and −0.23 in females in the right frontal region (F4) (*P=*0.04, Wilcoxon rank-sum test). The corresponding male and female differences in the right and left central regions (C3, C4) of the brain, though evident in the medians, were not statistically significant (*P=*0.4 and 0.4, respectively).

Median spindle density in both male and female diabetes patients differed little across the four brain regions; whereas median spindle density in male and female control subjects showed greater heterogeneity across those regions. Among both diabetes patients and controls, females tended to have higher median spindle density than males, with female-male differences especially large among controls (Table 2), consistent with literature reports (***Purcell et al. 2017***).

*Age*. Using linear regression analysis, we found little evidence that the patient-minus-control difference in spindle density exhibited a trend with age in any of the four brain regions (*P=*0.3, 0.5, 0.9, and 0.5 in regions F3, F4, C3, and C4, respectively) (Supplemental Table 4, Supplemental Figure 4).

### Spindle density and prediabetes

We were interested in seeing whether the presence of prediabetes, as a precursor to diabetes, in patients was also correlated with lower spindle density than matched controls. Comparison of 376 patients 18 yrs or older with a diagnosis of prediabetes and 376 age- and sex-matched control subjects showed evidence of lower spindle density among prediabetes patients in all four brain regions (Table 2). The medians of the prediabetes minus control differences were −0.09, −0.14, −0.12, and −0.15 spindles/min (*P=*0.04, 0.01, 0.03, and 0.03 in regions F3, F4, C3, and C4, respectively, Wilcoxon signed-rank test).

### External data validation

The SHHS recording montage consisted of electroencephalograms for the central regions (C3 and C4), but not the frontal regions (F3 and F4) of the brain. For both the C3 and C4 regions, subjects with self-reported diabetes had significantly lower spindle density (median=0.64 and 0.65 spindles/min) compared to the matched subjects without self-reported diabetes (median=0.87 and 0.85 spindles/min) (*P*=0.002 and 0.0007 in regions C3 and C4, respectively, Wilcoxon signed-rank test) (Table 2).

## DISCUSSION

Diabetes mellitus has been and remains a major public health issue in the US and elsewhere. The disease affects all organs because of high glucose levels in circulating blood and has many noticeable complications including coronary heart disease, chronic kidney disease, neuropathy, and stroke (***Collaborators 2023; Tomic et al. 2022; van Dieren et al. 2010***). Diabetes patients also experience brain complications (***Moheet et al. 2015; Seaquist 2010; van Sloten et al. 2020***) which are difficult to quantify and detect. A recent magnetic resonance imaging (MRI) study of 416 eligible EDIC (Epidemiology of Diabetes Interventions and Complications) participants and 99 demographically similar adults without diabetes showed that those with type 1 diabetes had significant brain atrophy compared to controls, mainly in the bilateral thalamus and putamen areas (***Jacobson et al. 2022***).

Sleep is important for diabetes management (***Briancon-Marjollet et al. 2015; Cappuccio et al. 2010; Chen et al. 2023; Farabi et al. 2016; Henson et al. 2024; Noga et al. 2024; Schipper et al. 2021; Spiegel et al. 1999)***. Here, we showed for the first time (to our knowledge) that patients with diabetes mellitus had significantly lower spindle density than those with controls during the N2 stage of the NREM sleep. Specifically, patients with diabetes mellitus exhibited on average about half the spindle density during sleep as their age- and sex-matched control subjects. We validated our finding on an external dataset. Sleep spindles are generated primarily in the thalamus where significant brain atrophy has been documented among type 1 diabetes patients (***Jacobson et al. 2022***).

Our results indicated that patients with at least one of the four major complications (CAD, CKD, stroke, and neuropathy) had similar spindle density to patients who had none of them. On the other hand, patients with prediabetes, like those with diabetes mellitus, may also show signs of lower spindle density compared to age- and sex-matched controls. Those findings suggest that spindle loss in diabetes patients may be gradual and may precede the occurrence of major complications.

Sleep spindles are involved in many sleep-related neurological functions including memory consolidation (***Andrillon et al. 2011; Fernandez and Lüthi 2020; Peyrache and Seibt 2020; Purcell et al. 2017; Schönauer and Pöhlchen 2018***). They also play a role in sensing and regulation of peripheral blood glucose in humans through coupling with other distinct EEG oscillations (***Tasali et al. 2008; Tingley et al. 2021; R. Vallat et al. 2023***). Accordingly, lower spindle density might exert a negative effect on peripheral glucose level sensing and regulation by the brain. One hypothesis worth considering is that diabetes contributes to a feedback loop involving high blood glucose levels perhaps leading to atrophy of the thalamus that, in turn, reduces spindle generation thereby exacerbating dysregulation of blood glucose control (Figure 1).

This hypothesis prompts several important questions. Does high blood glucose cause or contribute directly to atrophy of the thalamus? Does diabetes cause lower spindle density and possibly affect aspects of brain function by that mechanism? Specifically, does diabetes impair the brain’s capacity to sense and regulate peripheral glucose levels? Can intensive diabetes treatment reverse /reduce sleep spindle loss?

Answering some of those questions in the affirmative would have important public health implications. The Diabetes Control and Complications Trial (DCCT) trials demonstrated that intensive diabetes treatment to control hemoglobin A1C level does significantly improve the quality of life of patients with type 1 diabetes and reverse and slow down diabetes complications (Nathan et al. 1993). We suggest that some of the beneficial effects of the intensive diabetes treatment may accrue because they contribute to higher spindle density during sleep.

The principal strength of our study is its relatively large sample size (N=1,214) of high-quality EEG data from overnight in-laboratory studies in a certified sleep laboratory setting. The patient population referred to the sleep laboratory spans both urban and rural areas and a variety of races and ethnicities. Our study complements recent findings on the role of brain waves in sensing and regulation of glucose levels (***Tasali et al. 2008; Tingley et al. 2021; R. Vallat et al. 2023***). and on atrophy of the thalamus of patients with type 1 diabetes (***Jacobson et al. 2022***). Our finding of the reduced spindle density in patients with diabetes mellitus was validated in an external dataset.

Limitations are primarily those associated with restrictions on the data available to us in the hospital records. The patients referred to the sleep lab for sleep apnea evaluation/treatment can be heterogenous, but we have only limited data on individual patient characteristics. The diabetes mellitus diagnosis was obtained from medical records, and diabetes type was not specified for all patients, though at least two-thirds of the patients were type 2. We did not have race/ethnicity information for individual patients. We cannot totally rule out the possibility that the observed differences in spindle density might be associated with diabetes comorbidities and/or medications. Although we validated our finding on an external dataset (SHHS), the size of difference in spindle density was smaller in the SHHS dataset than in our dataset. The main difference between the SHHS data and our data is that diabetes in SHHS was self-reported whereas diabetes in our data were based on medical record. It is known that self-report diabetes may underestimate the prevalence of diabetes by as much as 50% (***Harris et al. 1987***). Also, subjects in SHHS (median age=∼70 yrs) were older than ours (median age=∼60 yrs). Nonetheless, additional replication of our findings is warranted.

## Conclusions

We discovered that patients with diabetes mellitus had significantly lower spindle density during stage 2 of the NREM sleep. We speculate that the reduced spindle density may be associated with brain atrophy in the bilateral thalamus as seen in patients with type 1 diabetes mellitus. We hypothesize that high glucose levels in blood may be a cause of thalamus atrophy leading to lower generation of spindles during sleep which, in turn, results in impaired peripheral blood glucose sensing/regulation. Further investigation of our hypothesis should expand our understanding of the relationship between diabetes and the brain. Reduced neurological complications from diabetes that accompany intensive diabetes treatment may be due to improved glucose control removing barriers to spindle generation.

## Acknowledgements

We thank the NIEHS Office of Scientific Computing for computing support.

## Author Contributions

D.Y collaborated on writing the first draft and carrying out data analysis. A.T., YL, and M.S. carried out data analysis. A.M-R. contributed to discussion and reviewed the manuscript. Z.F. contributed to data collection, discussion and reviewed the manuscript. D.M.U. contributed to substantially to revising the content and editing the manuscript. L.L. conceived the study, carried out data analysis, collaborated on writing the first draft. All authors approved the final version of the manuscript.

## Data Availability

The polysomnography data can be requested from the corresponding author. An institutional data use agreement will be required.

## References

Ahtiluoto, S., et al. (2010), ‘Diabetes, Alzheimer disease, and vascular dementia: a population-based neuropathologic study’, Neurology, 75 (13), 1195–202.

Andrillon, T., et al. (2011), ‘Sleep spindles in humans: insights from intracranial EEG and unit recordings’, J Neurosci, 31 (49), 17821–34.

Antal, B., et al. (2022), ‘Type 2 diabetes mellitus accelerates brain aging and cognitive decline: Complementary findings from UK Biobank and meta-analyses’, Elife, 11.

Arnold, S. E., et al. (2018), ‘Brain insulin resistance in type 2 diabetes and Alzheimer disease: concepts and conundrums’, Nat Rev Neurol, 14 (3), 168–81.

Barbiellini Amidei, C., et al. (2021), ‘Association Between Age at Diabetes Onset and Subsequent Risk of Dementia’, JAMA, 325 (16), 1640–49.

Bastaki, Salim (2019), ‘Diabetes mellitus and its treatment’, International Journal of Diabetes and Metabolism, 13 (3), 111–34.

Biessels, G. J. and Despa, F. (2018), ‘Cognitive decline and dementia in diabetes mellitus: mechanisms and clinical implications’, Nat Rev Endocrinol, 14 (10), 591–604.

Biessels, G. J., et al. (2006), ‘Risk of dementia in diabetes mellitus: a systematic review’, Lancet Neurol, 5 (1), 64–74.

Briancon-Marjollet, A., et al. (2015), ‘The impact of sleep disorders on glucose metabolism: endocrine and molecular mechanisms’, Diabetol Metab Syndr, 7, 25.

Buzsáki, G. (2015), ‘Hippocampal sharp wave-ripple: A cognitive biomarker for episodic memory and planning’, Hippocampus, 25 (10), 1073–188.

Cappuccio, F. P., et al. (2010), ‘Quantity and quality of sleep and incidence of type 2 diabetes: a systematic review and meta-analysis’, Diabetes Care, 33 (2), 414–20.

Chen, D. M., et al. (2023), ‘Altered sleep architecture in diabetes and prediabetes: findings from the Baependi Heart Study’, medRxiv.

Collaborators, G. B. D. Diabetes (2023), ‘Global, regional, and national burden of diabetes from 1990 to 2021, with projections of prevalence to 2050: a systematic analysis for the Global Burden of Disease Study 2021’, Lancet, 402 (10397), 203–34.

Cox, R., et al. (2020), ‘Sharp Wave-Ripples in Human Amygdala and Their Coordination with Hippocampus during NREM Sleep’, Cereb Cortex Commun, 1 (1), tgaa051.

DeFronzo, R. A., et al. (2015), ‘Type 2 diabetes mellitus’, Nat Rev Dis Primers, 1, 15019. edition, IDF Diabetes Atlas 2021 - 10th (2021), ‘IDF Diabetes Atlas’.

Eschenko, O., et al. (2008), ‘Sustained increase in hippocampal sharp-wave ripple activity during slow-wave sleep after learning’, Learn Mem, 15 (4), 222–8.

Farabi, S. S., Carley, D. W., and Quinn, L. (2016), ‘EEG power and glucose fluctuations are coupled during sleep in young adults with type 1 diabetes’, Clin Neurophysiol, 127 (8), 2739–46.

Fernandez, Laura M. J. and Lüthi, Anita (2020), ‘Sleep Spindles: Mechanisms and Functions’, Physiological Reviews, 100 (2), 805–68.

Ferrarelli, F., et al. (2007), ‘Reduced sleep spindle activity in schizophrenia patients’, Am J Psychiatry, 164 (3), 483–92.

Geva-Sagiv, Maya, et al. (2023), ‘Augmenting hippocampal–prefrontal neuronal synchrony during sleep enhances memory consolidation in humans’, Nature Neuroscience, 26 (6), 1100–10.

Gramfort, A., et al. (2013), ‘MEG and EEG data analysis with MNE-Python’, Front Neurosci, 7, 267.

Harris, M. I., et al. (1987), ‘Prevalence of diabetes and impaired glucose tolerance and plasma glucose levels in U.S. population aged 20-74 yr’, Diabetes, 36 (4), 523–34.

Helfrich, R. F., et al. (2019), ‘Bidirectional prefrontal-hippocampal dynamics organize information transfer during sleep in humans’, Nat Commun, 10 (1), 3572.

Henson, J., et al. (2024), ‘Waking Up to the Importance of Sleep in Type 2 Diabetes Management: A Narrative Review’, Diabetes Care, 47 (3), 331–43.

Jacobson, A. M., et al. (2022), ‘Brain Structure Among Middle-aged and Older Adults With Longstanding Type 1 Diabetes in the DCCT/EDIC Study’, Diabetes Care, 45 (8), 1779–87.

Joechner, A. K., et al. (2023), ‘Sleep spindle maturity promotes slow oscillation-spindle coupling across child and adolescent development’, Elife, 12.

Johnson, Jennifer M., Curtis, Ffion, and Durrant, Simon J. (2022), ‘Characterising the relationship between sleep stages and associated spectral power in diabetes’, Sleep Epidemiology, 2, 100048.

Karimi Abadchi, J., et al. (2020), ‘Spatiotemporal patterns of neocortical activity around hippocampal sharp-wave ripples’, eLife, 9, e51972.

Katsarou, A., et al. (2017), ‘Type 1 diabetes mellitus’, Nat Rev Dis Primers, 3, 17016.

Kellar, D. and Craft, S. (2020), ‘Brain insulin resistance in Alzheimer’s disease and related disorders: mechanisms and therapeutic approaches’, Lancet Neurol, 19 (9), 758–66.

Kodl, C. T. and Seaquist, E. R. (2008), ‘Cognitive dysfunction and diabetes mellitus’, Endocr Rev, 29 (4), 494–511.

Moheet, A., Mangia, S., and Seaquist, E. R. (2015), ‘Impact of diabetes on cognitive function and brain structure’, Ann N Y Acad Sci, 1353, 60–71.

Mölle, M., et al. (2006), ‘Hippocampal sharp wave-ripples linked to slow oscillations in rat slow-wave sleep’, J Neurophysiol, 96 (1), 62–70.

Moreno-Gonzalez, I., et al. (2017), ‘Molecular interaction between type 2 diabetes and Alzheimer’s disease through cross-seeding of protein misfolding’, Mol Psychiatry, 22 (9), 1327–34.

Nathan, D. M., et al. (1993), ‘The effect of intensive treatment of diabetes on the development and progression of long-term complications in insulin-dependent diabetes mellitus’, N Engl J Med, 329 (14), 977–86.

Ngo, H. V., Fell, J., and Staresina, B. (2020), ‘Sleep spindles mediate hippocampal-neocortical coupling during long-duration ripples’, Elife, 9.

Noga, D. A., et al. (2024), ‘Habitual Short Sleep Duration, Diet, and Development of Type 2 Diabetes in Adults’, JAMA Netw Open, 7 (3), e241147.

Olokoba, A. B., Obateru, O. A., and Olokoba, L. B. (2012), ‘Type 2 diabetes mellitus: a review of current trends’, Oman Med J, 27 (4), 269–73.

Ott, A., et al. (1999), ‘Diabetes mellitus and the risk of dementia: The Rotterdam Study’, Neurology, 53 (9), 1937–42.

Oyanedel, C. N., et al. (2020), ‘Temporal associations between sleep slow oscillations, spindles and ripples’, Eur J Neurosci, 52 (12), 4762–78.

Peyrache, Adrien and Seibt, Julie (2020), ‘A mechanism for learning with sleep spindles’, Philosophical Transactions of the Royal Society B: Biological Sciences, 375 (1799), 20190230.

Prerau, M. J., et al. (2017), ‘Sleep Neurophysiological Dynamics Through the Lens of Multitaper Spectral Analysis’, Physiology (Bethesda), 32 (1), 60–92.

Purcell, S. M., et al. (2017), ‘Characterizing sleep spindles in 11,630 individuals from the National Sleep Research Resource’, Nature Communications, 8 (1), 15930.

Quan, S. F., et al. (1997), ‘The Sleep Heart Health Study: design, rationale, and methods’, Sleep, 20 (12), 1077–85.

Reed, J., Bain, S., and Kanamarlapudi, V. (2021), ‘A Review of Current Trends with Type 2 Diabetes Epidemiology, Aetiology, Pathogenesis, Treatments and Future Perspectives’, Diabetes Metab Syndr Obes, 14, 3567–602.

Schipper, S. B. J., et al. (2021), ‘Sleep disorders in people with type 2 diabetes and associated health outcomes: a review of the literature’, Diabetologia, 64 (11), 2367–77.

Schönauer, M. and Pöhlchen, D. (2018), ‘Sleep spindles’, Curr Biol, 28 (19), R1129–r30.

Seaquist, E. R. (2010), ‘The final frontier: how does diabetes affect the brain?’, Diabetes, 59 (1), 4–5.

Siapas, A. G. and Wilson, M. A. (1998), ‘Coordinated interactions between hippocampal ripples and cortical spindles during slow-wave sleep’, Neuron, 21 (5), 1123–8.

Sims-Robinson, C., et al. (2010), ‘How does diabetes accelerate Alzheimer disease pathology?’, Nat Rev Neurol, 6 (10), 551–9.

Spiegel, K., Leproult, R., and Van Cauter, E. (1999), ‘Impact of sleep debt on metabolic and endocrine function’, Lancet, 354 (9188), 1435–9.

Staresina, Bernhard P., et al. (2023), ‘How coupled slow oscillations, spindles and ripples coordinate neuronal processing and communication during human sleep’, Nature Neuroscience, 26 (8), 1429–37.

Talukder, A., et al. (2024), ‘Comparison of power spectra from overnight electroencephalography between patients with Down syndrome and matched control subjects’, J Sleep Res, e14187.

Tasali, E., et al. (2008), ‘Slow-wave sleep and the risk of type 2 diabetes in humans’, Proc Natl Acad Sci U S A, 105 (3), 1044–9.

Tingley, D., et al. (2021), ‘A metabolic function of the hippocampal sharp wave-ripple’, Nature, 597 (7874), 82–86.

Tomic, D., Shaw, J. E., and Magliano, D. J. (2022), ‘The burden and risks of emerging complications of diabetes mellitus’, Nat Rev Endocrinol, 18 (9), 525–39.

Vallat, R., Shah, V. D., and Walker, M. P. (2023), ‘Coordinated human sleeping brainwaves map peripheral body glucose homeostasis’, Cell Rep Med, 4 (7), 101100.

Vallat, Raphael and Walker, Matthew P. (2021), ‘An open-source, high-performance tool for automated sleep staging’, eLife, 10, e70092.

van Dieren, S., et al. (2010), ‘The global burden of diabetes and its complications: an emerging pandemic’, Eur J Cardiovasc Prev Rehabil, 17 Suppl 1, S3–8.

van Sloten, T. T., et al. (2020), ‘Cerebral microvascular complications of type 2 diabetes: stroke, cognitive dysfunction, and depression’, Lancet Diabetes Endocrinol, 8 (4), 325–36.

Zhang, Guo-Qiang, et al. (2018), ‘The National Sleep Research Resource: towards a sleep data commons’, Journal of the American Medical Informatics Association, 25 (10), 1351–58.

