## supplementary tables and figures for "Differences in sleep spindle wave density between patients with diabetes mellitus and matched controls: implications for sensing and regulation of peripheral blood glucose"

**Table S1**. Prevalence of use of prescribed medications and of common complications among diabetes patients and control subjects matched on sex and age.

| Diabetic medications | Diabetes (N = 1,214) | | Control (N = 1,214) | |
|---|---|---|---|---|
| | N | % | N | % |
| DPP-4[1] inhibitors | 48 | 3.9 | 1 | 0 |
| GLP-1[2] agonists | 306 | 25.2 | 15 | 1.2 |
| Insulin | 629 | 51.8 | 0 | 0 |
| Metformin | 689 | 56.7 | 57 | 4.6 |
| SGLT2[3] inhibitors | 175 | 14.4 | 7 | 0.5 |
| Sulfonylureas | 132 | 10.8 | 7 | 0.5 |
| TZDs[4] | 15 | 1.2 | 2 | 0.1 |
| Others[5] | 10 | 0.8 | 3 | 0.2 |
| Major complications | | | | |
| Coronary artery disease | 257 | 21.1 | 81 | 6.6 |
| Chronic kidney disease | 130 | 10.7 | 11 | 0.9 |
| Neuropathy | 217 | 17.8 | 41 | 3.3 |
| Stroke | 61 | 5.0 | 4 | 0.3 |
| No listed complications | 711 | 58.5 | 1,086 | 89.4 |

[1] dipeptidyl peptidase IV
[2] glucagon-like peptide 1
[3] sodium-glucose transport protein 2
[4] thiazolidinediones
[5] alpha-glucosidase inhibitors, bile acid sequestrants, dopamine-2 agonists, meglitinides

**Table S2**. Comparison of selected sleep architecture characteristics between patients with diabetes and controls among those who underwent diagnostic PSGs.

| Group | N | Min. | 1st Qu. | Median | Mean | 3rd Qu. | Max. | *P* value[1] |
|---|---|---|---|---|---|---|---|---|
| | | Total sleep time (min.) | | | | | | |
| Diabetes | | 111.5 | 306.5 | 357.5 | 350.8 | 405.8 | 594.0 | |
| Control | 1,188 | 103.0 | 314.0 | 365.0 | 359.9 | 409.8 | 654.0 | 0.009 |
| Diabetes-Control | | -390.5 | -77.0 | -7.5 | -9.2 | 59.5 | 394.5 | |
| | | Sleep efficiency (%) | | | | | | |
| Diabetes | | 20.7 | 68.4 | 79.2 | 76.2 | 87.4 | 99.0 | |
| Control | 1,210 | 22.0 | 70.1 | 80.2 | 77.3 | 87.9 | 98.5 | 0.08 |
| Diabetes-Control | | -60.9 | -12.8 | -0.7 | -1.1 | 10.9 | 57.0 | |
| | | Sleep latency (min.) | | | | | | |
| Diabetes | | 0.0 | 5.3 | 13.3 | 23.2 | 28.6 | 253.9 | |
| Control | 1,210 | 0.0 | 6.3 | 14.4 | 21.8 | 27.7 | 196.7 | 0.9 |
| Diabetes-Control | | -169.8 | -14.0 | -1.1 | 1.5 | 14.6 | 250.1 | |
| | | N2 duration (min. × 2) | | | | | | |
| Diabetes | | 8.5 | 164.1 | 211.0 | 213.2 | 261.0 | 463.0 | |
| Control | 1,214 | 30.0 | 167.6 | 214.5 | 211.3 | 253.5 | 463.5 | 0.5 |
| Diabetes-Control | | -301.0 | -60.9 | 1.0 | 2.0 | 64.4 | 307.0 | |
| | | AHI (events/hour) | | | | | | |
| Diabetes | | 0.0 | 5.4 | 11.2 | 16.2 | 18.9 | 90.1 | |
| Control | 378 | 0.0 | 5.3 | 9.2 | 12.8 | 15.9 | 71.9 | 0.009 |
| Diabetes-Control | | -57.6 | -6.8 | 1.4 | 3.4 | 10.0 | 88,8 | |

[1] *P* value from two-sided Wilcoxon signed-rank test comparing diabetes patients to controls subjects; it tests the null hypothesis that the sleep variable specified is the same for diabetes patients and control subjects who underwent diagnostic PSGs.

**Table S3**. Comparison of six spindle characteristics between patients with diabetes mellitus and control subjects matched on sex and age. Estimates and test based on 1,214 matched patient-control pairs.

| Spindle Characteristic | Brain Region | Diabetes | | Control | | Paired difference | | |
|---|---|---|---|---|---|---|---|---|
| | | Median | IQR | Median | IQR | Median | IQR | *P* value[1] |
| Duration (sec) | Frontal (F3) | 0.75 | 0.10 | 0.76 | 0.09 | -0.01 | 0.14 | 0.9 |
| | Frontal (F4) | 0.74 | 0.10 | 0.76 | 0.09 | -0.01 | 0.14 | 0.9 |
| | Central (C3) | 0.76 | 0.10 | 0.78 | 0.09 | -0.02 | 0.14 | 0.9 |
| | Central (C4) | 0.76 | 0.09 | 0.78 | 0.09 | -0.02 | 0.14 | 0.9 |
| Absolute Power ($\log_{10} \mu V^2$) | Frontal (F3) | -0.67 | 0.29 | -0.59 | 0.24 | -0.09 | 0.38 | 0.8 |
| | Frontal (F4) | -0.64 | 0.28 | -0.57 | 0.23 | -0.07 | 0.37 | 0.9 |
| | Central (C3) | -0.63 | 0.25 | -0.56 | 0.21 | -0.08 | 0.35 | 0.8 |
| | Central (C4) | -0.62 | 0.24 | -0.55 | 0.21 | -0.07 | 0.33 | 0.9 |
| Relative Power | Frontal (F3) | 0.30 | 0.04 | 0.32 | 0.04 | -0.01 | 0.05 | 0.9 |
| | Frontal (F4) | 0.30 | 0.04 | 0.31 | 0.04 | -0.01 | 0.05 | 0.8 |
| | Central (C3) | 0.31 | 0.04 | 0.32 | 0.04 | -0.01 | 0.05 | 0.9 |
| | Central (C4) | 0.30 | 0.04 | 0.32 | 0.04 | -0.01 | 0.05 | 0.9 |
| Frequency (Hz) | Frontal (F3) | 12.25 | 0.55 | 12.28 | 0.50 | -0.03 | 0.73 | 0.8 |
| | Frontal (F4) | 12.26 | 0.53 | 12.29 | 0.49 | -0.04 | 0.73 | 0.8 |
| | Central (C3) | 12.62 | 0.63 | 12.70 | 0.63 | -0.08 | 0.81 | 0.7 |
| | Central (C4) | 12.61 | 0.58 | 12.69 | 0.63 | -0.06 | 0.78 | 0.9 |
| Number of Oscillations | Frontal (F3) | 8.64 | 1.15 | 8.84 | 1.05 | -0.22 | 1.56 | 0.9 |
| | Frontal (F4) | 8.60 | 1.13 | 8.84 | 1.10 | -0.24 | 1.63 | 0.9 |
| | Central (C3) | 8.93 | 1.15 | 9.17 | 1.10 | -0.25 | 1.64 | 0.9 |
| | Central (C4) | 8.87 | 1.12 | 9.12 | 1.12 | -0.28 | 1.60 | 0.8 |
| Symmetry | Frontal (F3) | 0.50 | 0.04 | 0.50 | 0.03 | 1.71e-3 | 0.06 | 0.3 |
| | Frontal (F4) | 0.50 | 0.05 | 0.50 | 0.03 | 1.30e-5 | 0.06 | 0.4 |
| | Central (C3) | 0.50 | 0.03 | 0.49 | 0.03 | 1.81e-3 | 0.05 | 0.2 |
| | Central (C4) | 0.49 | 0.04 | 0.49 | 0.03 | 8.94e-4 | 0.05 | 0.1 |

[1] *P* value from two-sided Wilcoxon signed-rank test applied to paired differences; it tests the null hypothesis that spindle density is the same for diabetes patients and controls subjects.

**Table S4**. Linear regression analysis of the difference in spindle density (spindles per min) between patients with diabetes and age-and sex-matched control subjects versus average age (yrs) of the matched pair for each brain region separately. Estimates and test based on 1,214 matched patient-control pairs.

| EEG channel | Estimated slope | Standard Error | t value | *P* value |
|---|---|---|---|---|
| F3 | -0.0025 | 0.0027 | -0.94 | 0.3 |
| F4 | -0.0018 | 0.0027 | -0.68 | 0.5 |
| C3 | -0.0005 | 0.0030 | -0.18 | 0.9 |
| C4 | -0.0018 | 0.0028 | -0.64 | 0.5 |

### Supplementary Figures

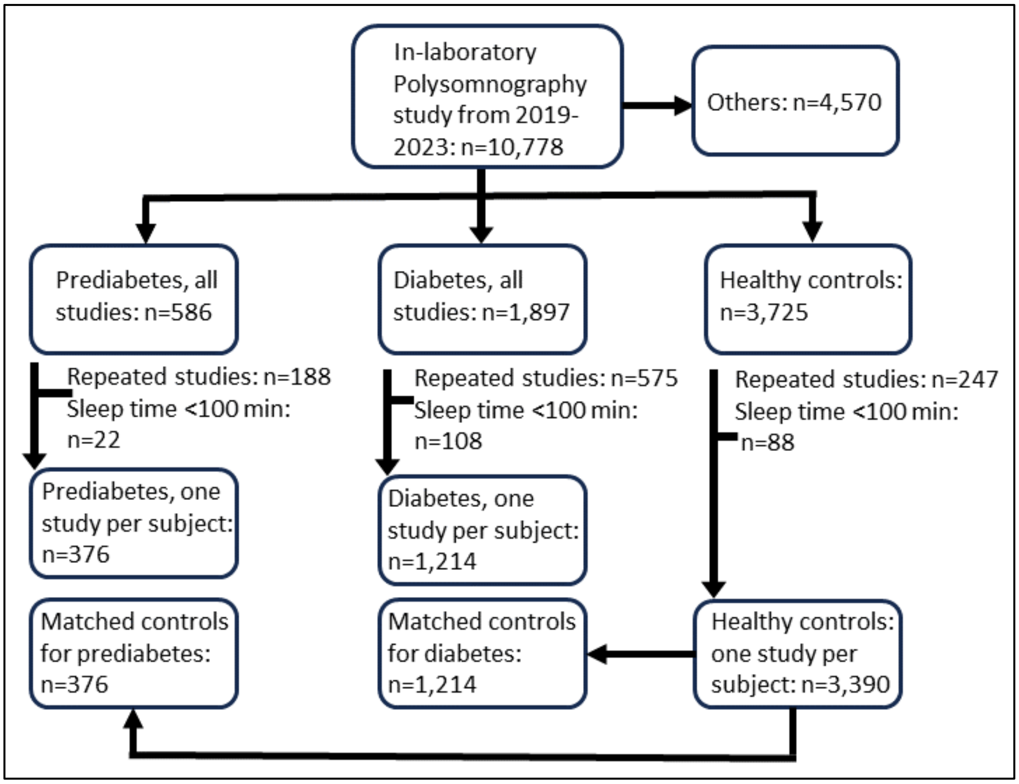

**Figure S1.** Patient inclusion flowchart.

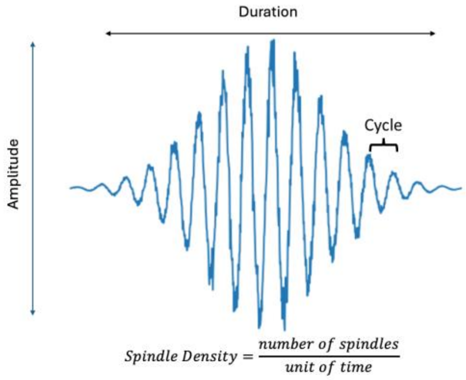

**Figure S2**. A cartoon illustration of a single spindle.

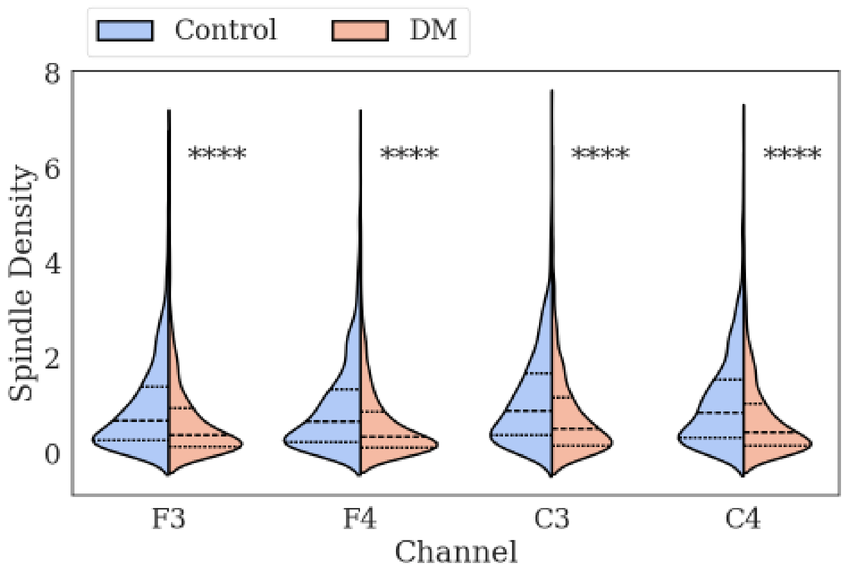

**Figure S3**. Differences in spindle density between patients with diabetes mellitus and controls. Violin plots (right, diabetes (DM); left, control) of the distribution of the spindle density in the frontal (F3 and F4) and central (C3 and C4) regions of the brain. Dashed lines show medians; dotted lines show the 1st and the 3rd quartiles. **** indicates $P<0.0001$ (two-sided Wilcoxon signed-rank test).

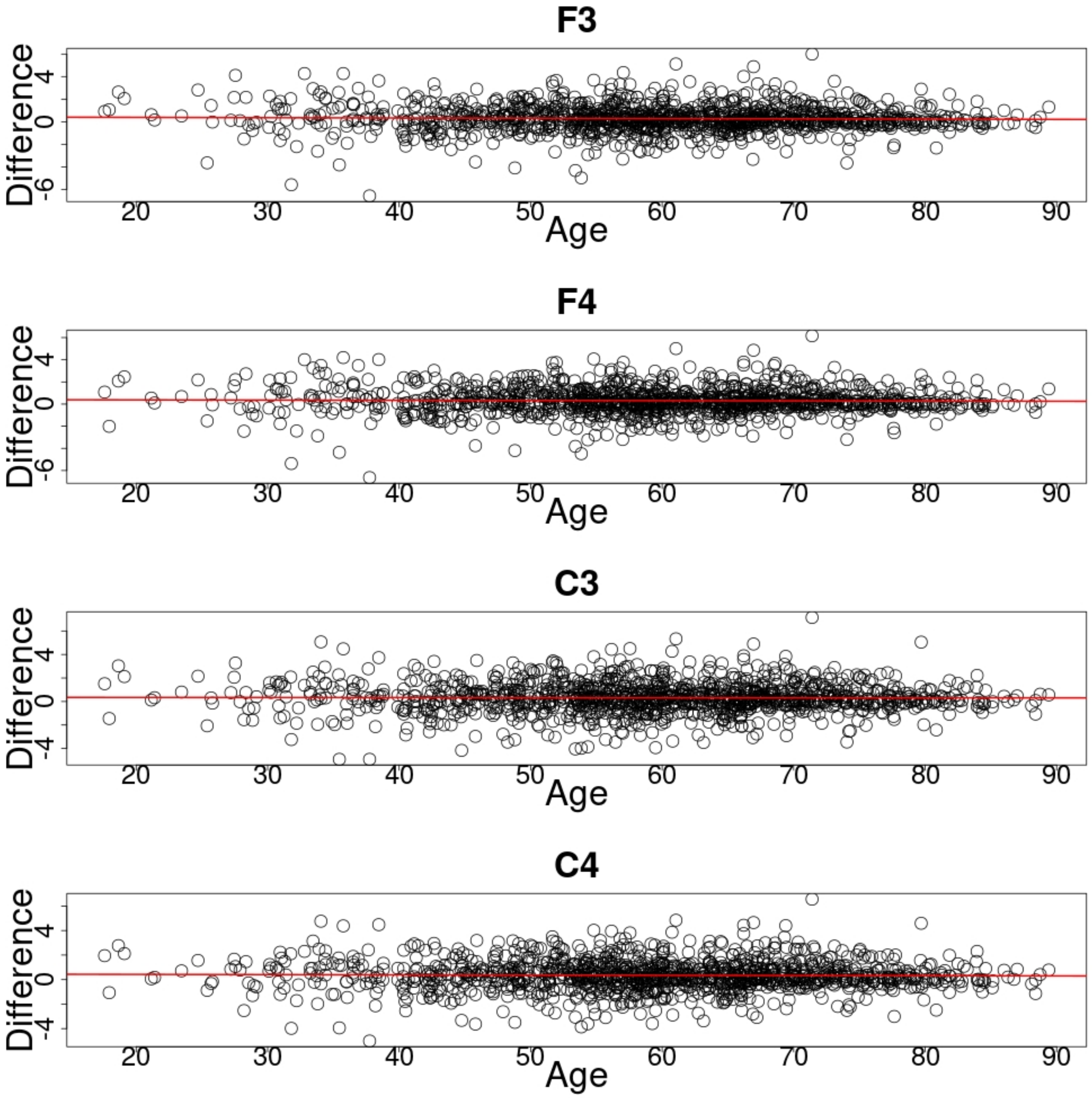

**Figure S4.** Linear regression analysis of the difference in spindle density (spindles per min) between patients with diabetes and age-and sex-matched control subjects versus average age (yrs) of the matched pair for each brain region separately. Plotted circles represent data for each of the 1,214 matched pairs. The red line is the fitted regression line (**Table S4)**.
